# Spherization of red blood cells and platelets margination in COPD patients

**DOI:** 10.1101/2020.04.03.20051748

**Authors:** Karim Zouaoui Boudjeltia, Christos Kotsalos, Daniel Ribeiro, Alexandre Rousseau, Christophe Lelubre, Olivier Sartenaer, Michael Piagnerelli, Jerôme Dohet-Eraly, Frank Dubois, Nicole Tasiaux, Bastien Chopard, Alain Van Meerhaeghe

**Affiliations:** Laboratory of Experimental Medicine (ULB 222), Medicine Faculty, Université libre de Bruxelles, CHU de Charleroi, Charleroi, Belgium; Computer science department, University of Geneva, Geneva, Switzerland; Internal Medicine, CHU de Charleroi - Hôpital Civil Marie Curie, Charleroi, Belgium; Microgravity Research Centre, Université libre de Bruxelles, Brussels, Belgium; Clinical Biology, Haematology Department, CHU de Charleroi, Charleroi, Belgium

## Abstract

**Rationale:** There are important interactions between Red Blood Cells (RBCs) and platelets in the bloodstream. These interactions lead to a phenomenon called margination. RBCs in pathological situations undergo biochemical and conformational changes leading to alterations in blood rheology.

**Aim:** RBCs shape in volunteers (21), stable (42) and exacerbated (31) COPD patients was analyzed. We studied the effect of the RBCs spherization on the platelets transport experimentally, *in vitro*, and by using numerical simulations.

**Methods:** RBC shape was estimated by the second moment of Pearson obtained through flow cytometry on *fsc* histogram. *In vitro* experiments were performed to analyze the effect of RBC shape on platelets adhesion/aggregation in dynamic conditions. Neuraminidase treatment was used to induce RBCs spherization. Numerical simulation were performed to determine the effect of RBCs spherization on platelets mean square displacement (MSD) to provide a physical explanation.

**Results:** Significant increase of RBC sphericity was observed in COPD patients compared to volunteers (Kruskal-Wallis: p<0.0001). *In vitro* experiments, at shear rate of 100 s^-1^, we observed that RBCs treated with neuraminidase mainly affect the number of platelet aggregates (p = 0.004). There was no change in the aggregates size. At a shear rate of 400 sec^-1^ neuraminidase treatment changes both the size of the aggregates (p = 0.009) and the number of platelet aggregates (p = 0.008).

Numerical simulations indicated that RBCs spherization induces an increase of MSD and the effect was more pronounced when the shear rate increased.

**Conclusion:** Our results show that the RBCs of COPD patients are more spherical than those of healthy volunteers. Experimentally we observe that the RBCs spherization induces an increase platelet transport to the wall. Additional studies are needed to better understand the possible association between the RBCs effect on the platelets transport and the increased cardiovascular events observed in COPD patients.

## Introduction

Chronic obstructive pulmonary disease (COPD) is an important cause of mortality worldwide and is associated with the development of cardiovascular events ^1,2^. Donaldson et al ^3^ reported that the risk of acute cardiovascular events increased during an exacerbation of chronic obstructive pulmonary disease (ECOPD). The risk of myocardial infarction 1 to 5 days after an ECOPD episode increased 2.3 fold (95% CI 1.1 to 4.7; p= 0.03) and that the risk of stroke 1 to 49 days after ECOPD increased 1.3 fold (95% CI 1.0 to 1.06; p=0.05). Another study showed that ECOPD patients died within 24h of hospitalization due to a cardiac failure and thromboembolism ^4^.

Chronic inflammation has been considered, as a determinant factor of multimorbidities in COPD patients. Extensive cross talk between inflammation and coagulation exists, and platelets, monocytes and their interaction play a pivotal role in chronic inflammation and development of cardiovascular diseases (CVD) ^5^. Activated platelets secrete numerous of substances such as chemokines, cytokines and molecules involved in haemostasis. Platelet degranulation leads to increased expression of P-selectin on platelet surface membrane, whereas a conformational change in the integrin αIIβ3 results in increase fibrinogen binding. Moreover, fibrinogen binding results in platelets aggregation, clot formation and adhesion of platelets to endothelial cells surface.

In light of the various published scientific works, we know that there are important interactions between Red Blood Cells (RBCs) and platelets in the bloodstream. These interactions lead, among other things, to a phenomenon called margination.

In blood flow, the distribution of platelet concentration is not uniform. Due to the phenomenon of margination, the concentration of the platelets to the wall increases, and conversely, the concentration of platelets in the center of the blood vessel decreases ^6,7^. This imbalance is interpreted from a finalist point of view as being a need for platelets at the vascular wall in case of injury to limit bleeding.

However, an increase in this margination process may also modified platelet-vascular wall interactions and lead to uncontrolled platelet aggregation formation and endothelial dysfunctional responses. This in response to possible changes in local viscosities that have not yet been clearly determined experimentally.

It is well known that RBCs in pathological situations undergo biochemical and conformational changes leading to alterations in blood rheology ^8^.

Spherization of RBCs has been demonstrated in sepsis but also in other pathologies inducing chronic or acute systemic inflammation ^9^.

In COPD patients some reports of constitutional changes in the composition of membrane lipids in RBCs exist, but no systematic study of the form of globules and its consequences has been performed.

In this work we propose an analysis of the shape of RBCs in stable and exacerbated COPD patients.

We will also analyze the effect of the spherization of red blood cells on margination of platelets experimentally and numerically.

## Material and methods

### Clinical data and blood parameters

This monocentric cross-sectional study enrolled 94 subjects between January 2018 and October 2019. Control subjects (n=21) had normal spirometry and no history of respiratory symptoms. COPD patients had an anamnesis compatible with the diagnosis, a smoking history of at least 10 pack-years, persistent airflow limitation on post-bronchodilator spirometry (Forced expiratory volume in 1 s/forced vital capacity (FEV_1.0_)/FVC ratio <0.70. (GOLD classification). Stable disease was defined as no exacerbation of COPD in the previous 8 weeks and no regular oral corticosteroids (n= 42). 40% of them were recruited during their follow-up consultation for oxygen therapy. Exacerbated COPD patients were recruited upon hospital admission (n=31). During acute COPD exacerbations, all patients received controlled oxygen therapy, nebulised bronchodilators and systemic corticosteroids for at least 5 days.

Exclusion criteria in both groups of COPD patients were asthma, bronchiectasis, pneumonia, pulmonary embolism, coronary artery disease, heart, hepatic and renal failure. No patient was receiving clopidogrel. Two stable COPD patients were treated for obstructive sleep apnea syndrome.

All subjects provided written informed consent. The Ethics Committee of the CHU-Charleroi approved the study (N°: OM008)

Serum samples were collected in vacuum tubes without anticoagulant. Plasma samples were harvested in citrated vacuum tubes, immediately processed. Whole blood was collected on EDTA-treated tubes. CRP, Urea and Creatinine were evaluated on ARCHITECT, Ci 4100 (ABBOTT®). Fibrinogen was determined by the Clauss method on a STA® automate (Stago). Blood cells were determined on a SYSMEX Hemato, XN-1000.

All the parameters were measured on the same day.

### RBC’s shape analysis

Data were collected on the Flow Cytometer Macs Quant® (Miltenyi Biotec). RBC samples were analyzed within 90 mins. The general principles of the measurement have been previously published by Piagnerelli et al ^9^.

### In vitro effect of Red Blood Cells spherization on platelet adhesion and aggregation

Blood samples were collected in 3.6 ml sodium citrate blood tubes and centrifuged at 1480 RPM for 5 minutes at room temperature. The supernatant was discarded and the RBCs’ fraction was transferred to a 15 ml tube. RBCs were washed by three consecutive steps of adjusting the tube’s total volume with HBSS, centrifugation for 5 min. at 1480 RPM and the removal of the supernatant. The RBCs’ fraction was then resuspended to the initial volume using RPMI (3.6ml).

Finally, the RBC solution was divided by two sampling tubes. One treated with 25mU Neuraminidase in RPMI (conditions 1) and the other with equivalent volume of RPMI (treatment control). The tubes were incubated at 37°C overnight.

After overnight incubation, tubes were centrifuged at 1480 RPM for 5 minutes and the supernatant discarded and replaced by the platelet rich plasma (PRP) portion obtained by the centrifugation at 950 RPM for 10 min of fresh blood samples from the same subjects drawn by venipuncture into two sodium citrate tubes of 3.6 ml. The final concentration of RBC in each test condition was controlled by flow cytometry.

Using the Impact-R platelet function analyzer, platelet aggregates formation was induced by placing 130 μl of the different RBC solutions (control and treatment) on a polystyrene well and subjecting it to different flows (100 s^-1^ and 400 s^-1^) conditions using disposable Teflon conical rotors. The method has been previously published ^7^.

### Numerical analysis of Red Blood Cells spherization on platelet margination process

The numerical counterpart of impact-R experiments is conducted using a high-performance computing software for ab initio modelling of cellular blood flow ^10^. The framework models blood cells individually (trajectories), like red blood cells (RBCs) and platelets, including their detailed non-linear viscoelastic behavior and the complex interaction between them. Our goal is to quantify the impact of RBC spherization on platelet transport and use this numerical tool to complement the experiments conducted using the Impact-R platelet analyzer.

The computational tool is based on three modules, namely the fluid solver, the solid solver and the fluid-solid interaction (FSI). The fluid solver (blood plasma resolution) is based on the lattice Boltzmann method (LBM) ^11^ and the module uses Palabos as for the implementation of the LBM. Palabos (https://palabos.unige.ch/) ^12,13^ stands for Parallel Lattice Boltzmann Solver and is an open source Computational Fluid Dynamics (CFD) library. The FSI (coupling of the two phases) is done via an immersed boundary method known as multidirect forcing scheme ^14^, implemented in Palabos. Lastly, the solid solver (blood cells resolution) is based on the nodal projective finite elements method (npFEM) ^15^ and the module uses the npFEM library developed and maintained by Palabos team. The framework is designed to operate on hybrid computational systems, i.e., fluid phase solved on CPUs (central processing units) while solid phase solved on GPUs (graphics processing units). The numerical experiments are performed on Piz Daint at the Swiss National Supercomputing Center, which according to the list TOP500 (https://www.top500.org/) (November 2019) is ranked 6^th^ worldwide and 1^st^ in Europe.

## Results

### Clinical data

Demographics characteristics are in table 1 and 2. The three groups were well matched with respect to age, gender and body mass index. None of the controls were smokers.

**Table 1.** Demographics data

|  | **Control subjects**<br>**(group A)** | **Stable COPD**<br>**(group B)** | **Exacerbation COPD**<br>**(group C)** | **All** | **B vs C** |
| --- | --- | --- | --- | --- | --- |
| Participants, *n* | 21 | 42 | 31 |  |  |
| Age | 61(58–68) | 67.5(61-74) | 65(61.2-71.7) | NS |  |
| BMI (kg/m<sup>2</sup>) | 25.1(23.5-26.4) | 26(21.2-31) | 25(20.2-31.6) | NS |  |
| Sex (male/female) | 12/9 | 16/26 | 16/15 | NS |  |
| Current smokers | 0% | 14% (6/42) | 26% (8/31) | NS |  |
| SABA_SAMA |  | 86% | 90% |  | NS |
| LAMA |  | 5% | 13% |  | NS |
| LABA |  | 2.5% | 3.2% |  | NS |
| LABA_LAMA |  | 17% (7/42) | 38.7% (12/31) |  | 0.03 |
| LABA_ICS |  | 7% (3/42) | 3.2%(1/31) |  | NS |
| LABA_LAMA_ICS |  | 67% (28/42) | 42% (13/31) |  | 0.03 |
| ICS |  | 0% | 0% |  | NS |
| Oxygen therapy |  | 40% | 25.8% |  | NS |
| ASA | 14% (3/21) | 55% (23/42) | 55% (17/31) | 0.004 |  |
| Statins | 4.7% (1/21) | 36% (15/42) | 25.8% (8/31) | 0.02 |  |
| Type 2 Diabetes | 0% | 19% (8/42) | 16.1% (5/31) | 0.03 |  |
| Hypertension | 19%(4/21) | 19% (8/42) | 19.3% (6/31) | NS |  |
| Renal insufficiency | 0% | 0% | 0% | NS |  |

**Table 2.** Respiratory function

|  | **Stable COPD**<br>**(group B)** | **COPD**<br>**Exacerbation**<br>**(group C)** | **P Value** |
| --- | --- | --- | --- |
| *n* | 42 | 31 |  |
| FEV1.0_FVC_Ratio % | 44.5(35-55) | 51.6(41.5-59) | NS |
| FEV1.0_%Predicted | 28.9(24-39) | 35(27.2-44.7) | NS |
| FEV1.0_ml | 770(540-962) | 851(652-1187) | NS |
| Gold II A | 2 |  |  |
| Gold II B | 4 |  |  |
| Gold II C | 2 |  |  |
| Gold II D |  | 4 |  |
| Gold III B | 10 |  |  |
| Gold III D |  | 15 |  |
| Gold IV B | 14 |  |  |
| Gold IV D | 10 | 12 |  |

Median (25%-75%), Mann-Whitney test
FEV<sub>1.0</sub>: forced expiratory volume in 1s; FVC: forced vital capacity.

At the level of the maintenance medications in the COPD populations, the only differences were in the greater number of patients under triple therapy in stable COPD, and the greater number under bi therapy (LABA-LAMA) in the exacerbated COPD patients. Statins (n=1) and acetylsalicylic acid (n=3) were rarely prescribed in control subjects.

Patients with COPD had moderate to very severe airflow limitation (GOLD stages 2-4) with a median value of FEV_1.0_ of 770 ml in the stable group and 851 ml in the exacerbated group. Blood parameters table 3.

**Table 3:** Blood parameters

|  | Control subjects<br>(group A) | Stable COPD<br>(group B) | COPD<br>Exacerbation<br>(group C) | ANOVA<br>on Rank |
| --- | --- | --- | --- | --- |
| Hct % | 44.2(42.7-46.8) | 43.2(39.8-45.6)* | 40.6(36.3-42.6)* † | <0.001 |
| Hb (g/dl) | 15(14.5-15.7) | 14.1(13.1-15.2) | 12.8(11.8-14)* † | <0.001 |
| RBC (x 10<sup>6</sup>/mm<sup>3</sup>) | 4.97(4.8-5.23) | 4.75(4.37-5.11) | 4.4(4.04-4.68)* † | <0.001 |
| MCV (fl) | 89.8(85.9-92.5) | 91.4(87.7-96.8) | 92.3(89-95.2) | 0.08 |
| RDW % | 13(12.6-13.3) | 13.5(13.1-14.2)* | 14.05(13.3-15.2)* | <0.001 |
| Plt (x 10<sup>3</sup>/mm<sup>3</sup>) | 247(204-292) | 265(219-307) | 265(222-327) | 0.74 |
| MPV (fl) | 10.4(9.8-11.5) | 10.6(9.9-11.2) | 10.1(9.9-10.7) | 0.016 |
| WBC (10<sup>3</sup>/mm<sup>3</sup>) | 6.74(5.31-7.66) | 8.59(7.28-10.15)* | 10.12(8.42-14.6)* | <0.001 |
| Neutro (10<sup>3</sup>/mm<sup>3</sup>) | 3.54(2.6-4.29) | 5.62(4.44-7.1)* | 8.46(5.67-11.68)* † | <0.001 |
| Lymp (10<sup>3</sup>/mm<sup>3</sup>) | 2.03(1.72-2.99) | 1.76(1.27-2.33) | 1.36(0.81-1.85)* † | <0.001 |
| Mono (/mm<sup>3</sup>) | 530(465-610) | 650(490-780) | 725(360-897) | 0.19 |
| Fibrinogen (g/l) | 3.27(2.72-3.75) | 4.12(3.4-4.85)* | 4.85(3.66-6.55)* | <0.001 |
| CRP (mg/l) | 1(1-2) | 3(1-6) | 21.5(3.7-52.5)* † | <0.001 |
| Urea (mg/dl) | 35.5(32.5-41.7) | 32(24.7-38.5) | 35(27.2-43.7) | 0.24 |
| Creat (mg/dl) | 0.85(0.81-1.01) | 0.77(0.66-0.95) | 0.78(0.72-0.89) | 0.09 |

Median (25%-75%), \* < 0.05 vs control, †<0.05 vs Stable COPD

### Red Blood Cells sphericity

Figure 1 shows the sphericity of RBCs estimated by the second moment of Pearson based on the *fsc* histogram obtained through flow cytometer ^9^. We observe a significant increase of RBCs sphericity in stable COPD patients - 0.46 (-0.54 - -0.4), exacerbated COPD patients - 0.54 (-0.54 - -0.32)compared to volunteers -0.57 (-0.71 - -0.54) (Krusal-Wallis: p<0.0001). No significant difference was observed between stable and exacerbated COPD patients.

**Figure 1:**
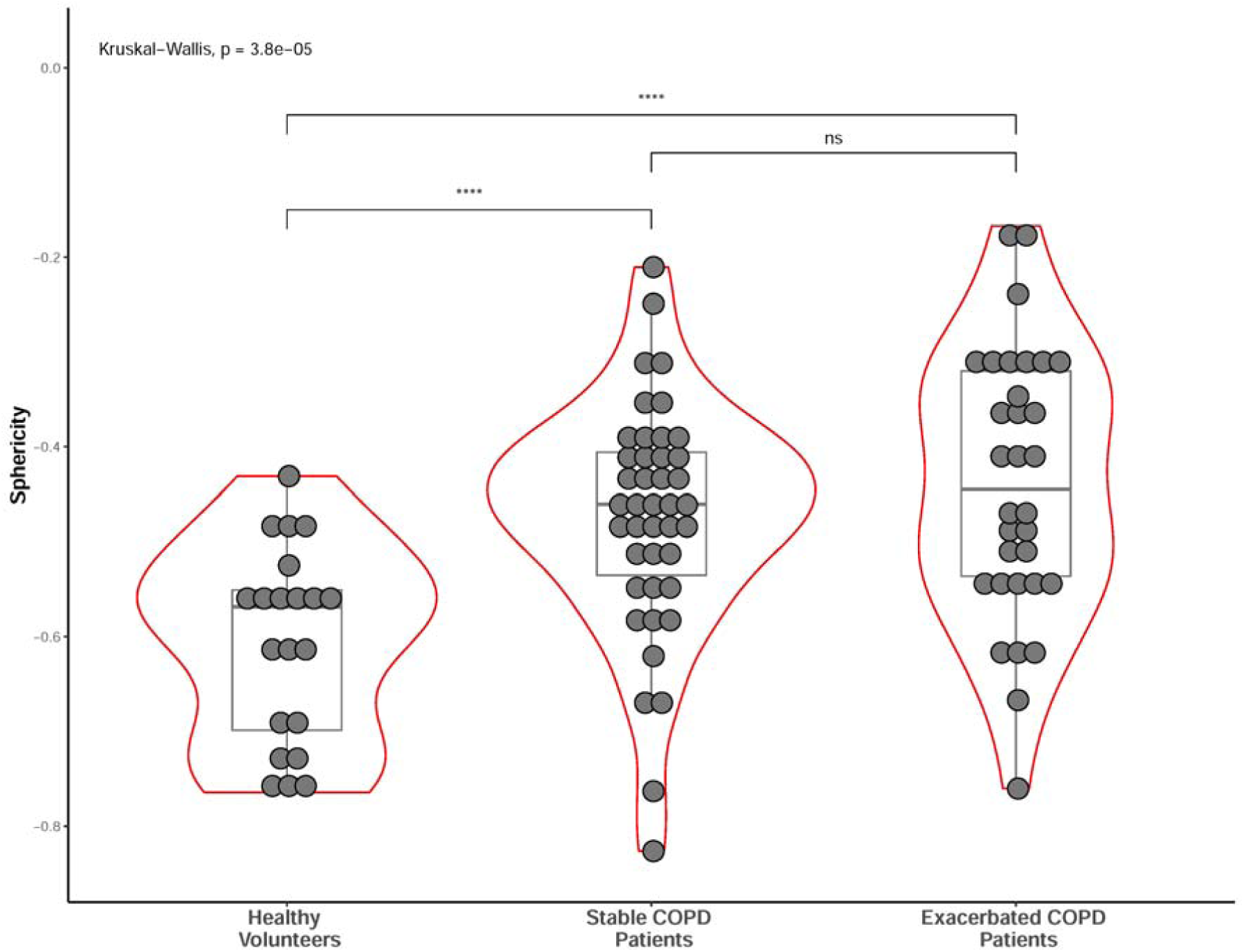
Red Blood Cells sphericity in volunteers and COPD patients.

### Effect of Red Blood Cells sphericity on platelets deposition and aggregation

The objective of this experiment was to analyze the effect of the shape changes of RBCs on platelet transport. To do this, we used washed RBCs from healthy volunteers (n = 7). Part of the RBC was kept as a control and the rest was treated with neuraminidase. Indeed, we have previously demonstrated in different works that the loss of sialic acid in different clinical and *in vitro* situations was associated with spherization of GRs.

After 24 hours of incubation with sialidase (0.025 U / ml), the RBCs were washed and added to freshly drawn platelet-rich plasma from the same volunteers. In this way only the shape of the red blood cells was changed. The sphericity, based on *fsc* histrogram, was -0.755 ± 0.12 vs -0.601 ± 0.16 for the neuraminidase treatment (p = 0.02).

Samples final hematocrit was 36% ± 1.6%.

Two shear rates were applied to mimic blood flow: 100 s^-1^ and 400 s^-1^.

At shear rate of 100 s^-1^ (Fig 2A) we observed that RBCs treated with neuraminidase mainly affects the number of platelet aggregates (p = 0.004). There was no change in the aggregates size.

**Figure 2:**
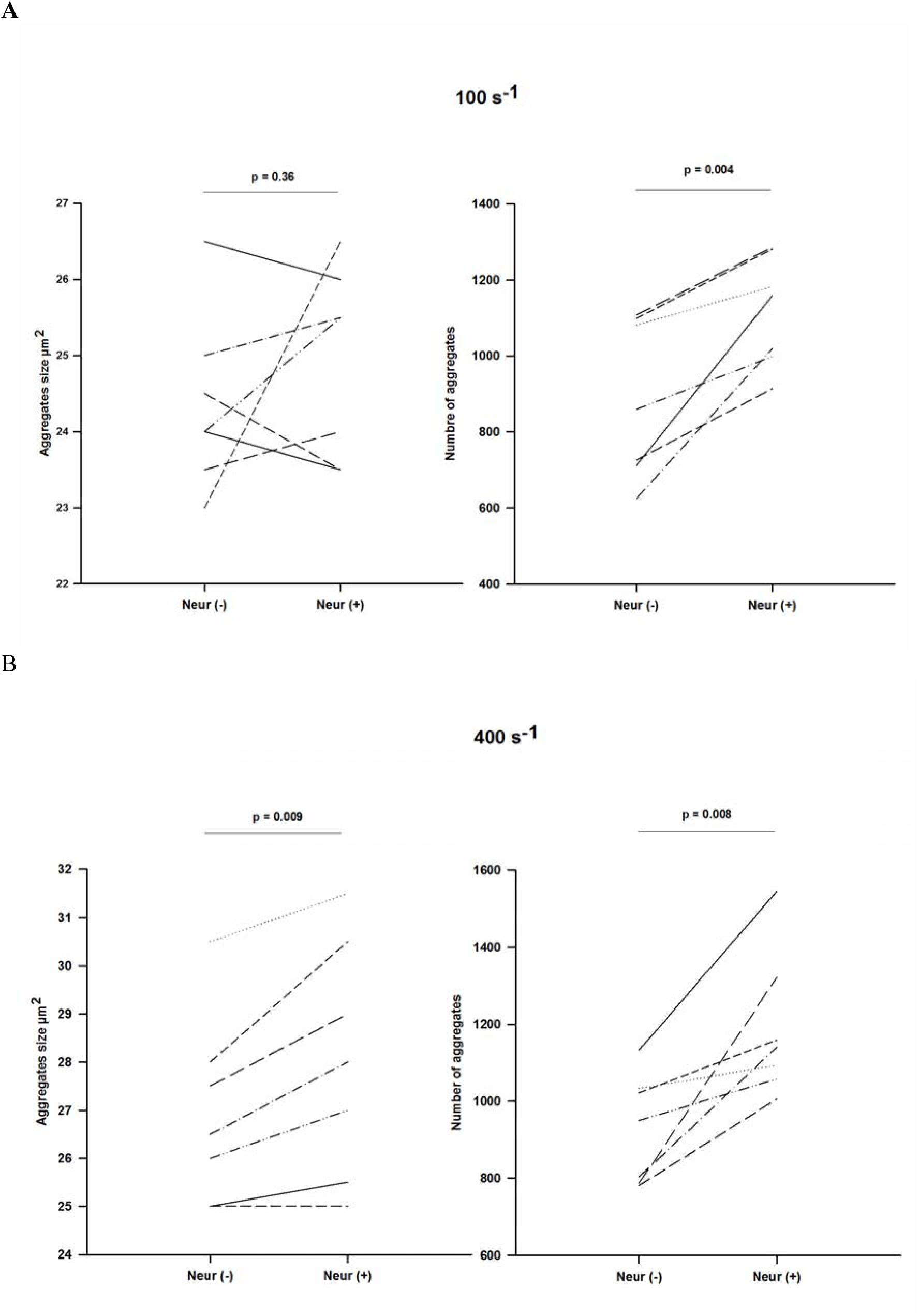
Impact-R results at shear rate 100 s^-1^ and 400 s^-1^ with control RBCs and neuraminidase treated RBCs.

At a shear rate of 400 sec^-1^ (Fig 2B) neuraminidase treatment changes both the size of the aggregates (p = 0.009) and the number of platelet aggregates (p = 0.008).

#### Numerical Simulations

The volume of interest in impact-R device spans the 1×1 mm^2^ observation window and has a 0.82 mm height. Given that a single mm^3^ of treated blood contains a few million RBCs and a few hundred thousand platelets, the computational cost of fully resolving this volume is still challenging, if not impossible with the current supercomputers and tools. For this reason, we focus on a much smaller region and simulate a cube of 50^3^ μm^3^ (**x** × **y** × **z**). The flow direction is parallel to the z-axis, periodic boundaries are applied in the x, z directions, and the y-axis is the wall-bounded direction. The upper wall is moving with such velocity, that the desired shear rate (100/400 s^-1^) is reached, while the bottom wall is fixed. The hematocrit is 35% in every numerical experiment and the domain is initialized by randomly positioning blood cells.

The platelets are simulated as nearly rigid oblate ellipsoids with diameter 3.6 μm, thickness 1.1 μm, and volume 6.8 fL (non-activated platelets). As for the shape of RBCs, three different shapes are considered, i.e., the normal biconcave and two spherized versions (v0 & v1) as seen in Figure 3. Every experiment uses just one type of RBC shape. The normal RBC shape is generated by producing points that satisfy the following equation

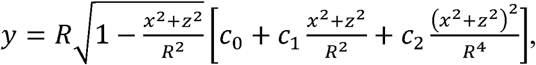

where (R, c_0_, c_1_, c_2_) = (3.91 μm, 0.1035805, 1.001279, -0.561381) ^16^. The area and volume of normal RBCs are approximately 133 μm^2^ and 93 μm^3^. The shape of spherized RBCs is generated by producing points that satisfy the following equation

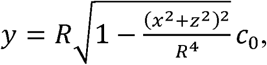

where (R, c_0_) = (3.5 μm, 0.4285714) for v0 and (R, c_0_) = (3.5 μm, 0.5714286) for v1. The area/volume of v0 are (108 μm^2^, 90 μm^3^) and for v1 are (123 μm^2^, 120 μm^3^). A remark at this point is that the material of the RBC remains the same for all experiments and a detailed presentation/validation of the material model can be found in ^10,15^. The sphericities of normal RBC/v0/v1 are 0.746/0.899/0.956. The change of sphericity in the numerical experiments is in accordance to the change observed in the in vitro experiments (∼30%).

**Figure 3:**
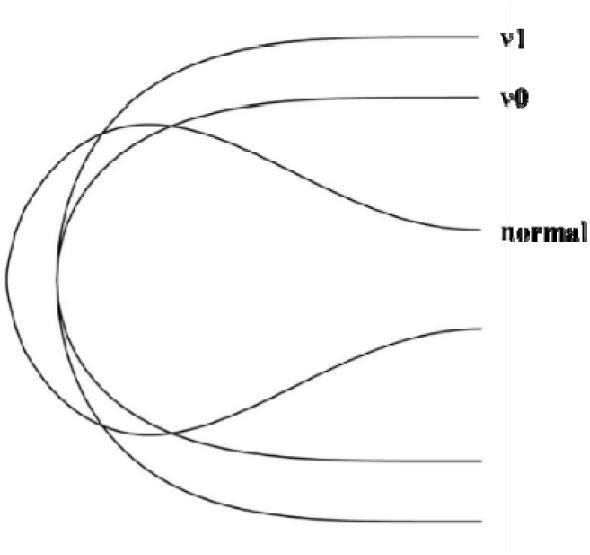
Different RBC shapes as generated by the equations (cross-sections). The 3D objects are produced by revolving the curves around the central axis.

Figure 4 presents snapshots from the simulations. Our goal is to quantify the effect of the RBC shape on platelet margination. We deploy the mean square displacement (MSD) in the wall-bounded direction. The MSD is defined as ⟨ (*y*_*i*_(*t*) –*y*_*i*_(*t*_0_))^2^⟩, with y_i_ the position of platelet i in the wall-bounded y-direction and ⟨■⟩ is the averaging over the platelets of interest. The averaging concerns either all available platelets, i.e., the ones in the RBC-rich layer (RBC-RL) and platelets in the Cell Free Layer (CFL), or only platelets in the RBC-RL. The MSD encodes how fast blood particles move away from their initial position, and thus the slope of MSD over time is traditionally linked with the diffusion coefficient of platelets. In order to map the effect of RBC shape on platelet transport, we consider the MSD in the RBC-RL, thus excluding the effects of the boundaries. We do not investigate the CFL properties, e.g., thickness, and for this reason we consult other studies ^17^, considering the thickness of the CFL to be one platelet diameter. From the fully resolved 3D simulations, we track at every time step the positions of platelets, and we sample the output at every 10 ms to produce further data like the MSD and the average distance of platelets from the walls. The metrics presented below are independent of the initialization of the cell field since they concern the collective behavior of platelets, i.e., any repetition of the numerical experiments leads to approximately the same results, given that the statistical sample is sufficient (number of platelets and resolved physical time).

**Figure 4:**
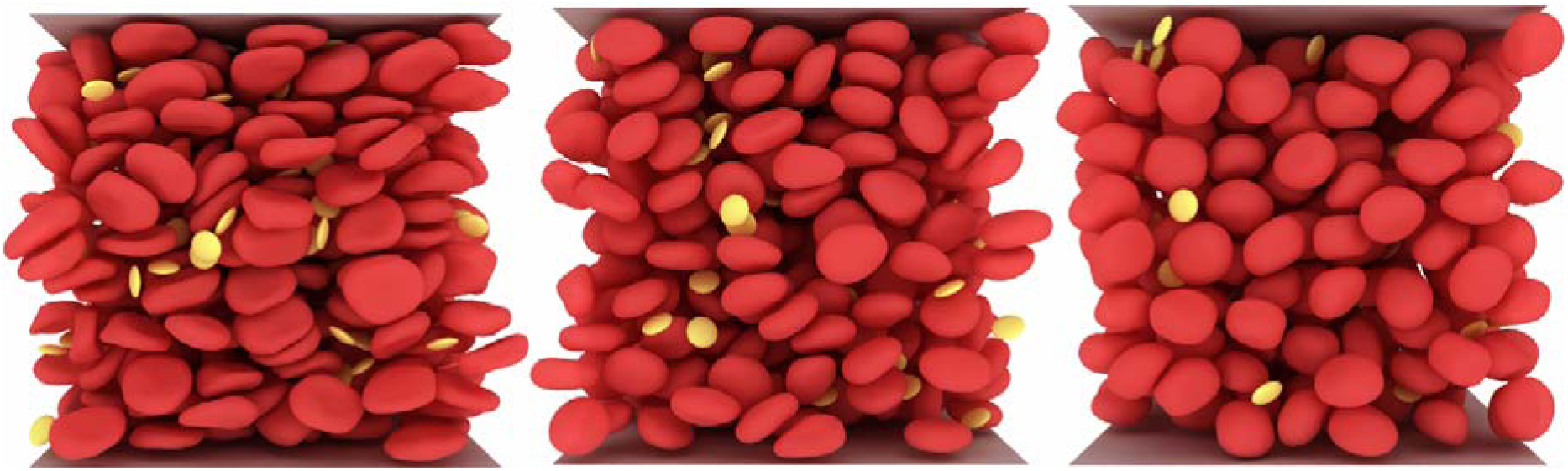
Shear flow generated by our computational framework (10) in a cubic domain of size 50^3^ μm3. From left to right, normally shaped RBCs, v0 spherized RBCs & v1 spherized RBCs. The hematocrit is 35% and the shear rate is either 100 or 400 s-1 according to the *in vitro* experiments.

Figure 5 presents the MSD evolution for a 500 ms time interval. The numerical experiments clearly suggest that the spherization of RBCs has a great impact on platelet margination, i.e., increased margination for more spherical shapes and higher shear rates. For shear rate 400 s^-1^, the v0 case gives a similar trend with the normal RBC shape. An explanation is that for 400 s^-1^ the deformations are much higher (compared to 100 s^-1^) and thus there is one more factor affecting the transport of platelets, i.e., deformability in addition to shape. A more spherical RBC is less deformable than a biconcave one, and as reported by Chang and al ^18^, less deformable RBCs lower the transport of platelets towards the vessel walls (shape factor has opposite effect from deformability). Figure 6 presents the normalized average distance of platelets from the walls over time. The numerical experiments consistently show that the platelets move towards the walls. Furthermore, for shear rate 400 s^-1^ the slopes are higher than the 100 s^-1^ suggesting that the margination is more pronounced in the former case. To summarize, Figure 5 and Figure 6 support the experimental findings and provide a consistent view on how the RBC shape affects the platelets transport.

**Figure 5:**
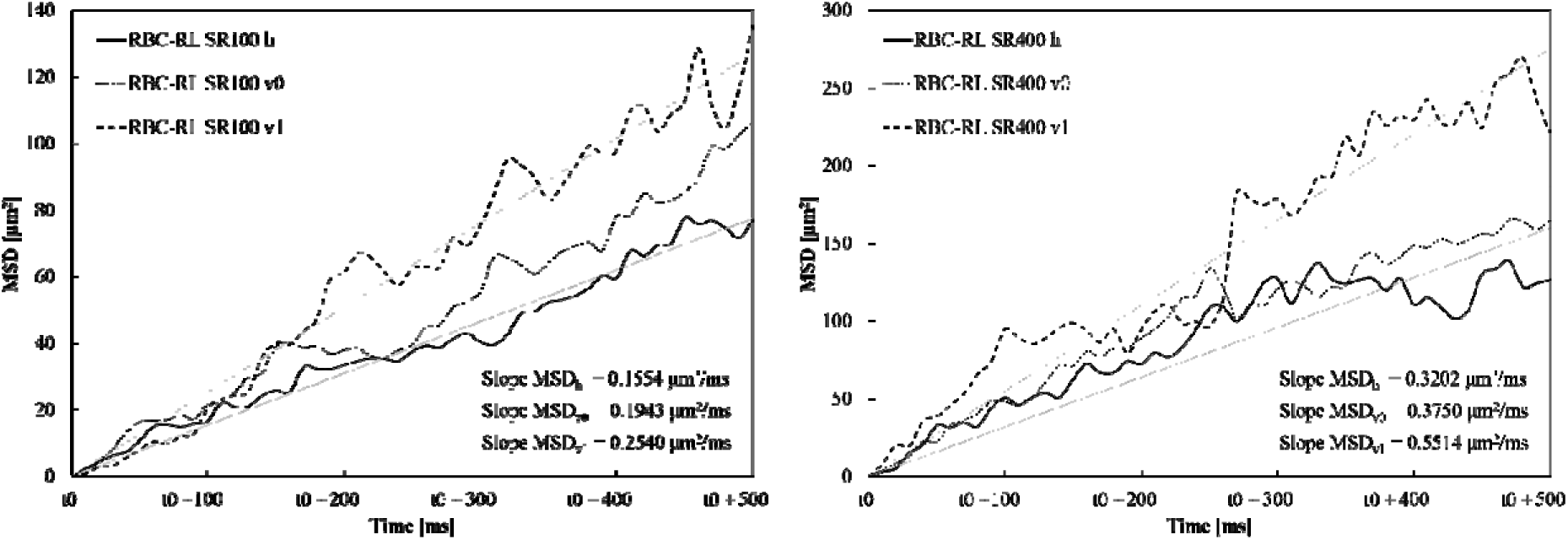
MSD with averaging over the platelets of the RBC-RL over time. The slopes at the bottom right of every image are obtained from a linear fitting (visible straight lines) on each curve. The t0 corresponds to 300 ms and 100 ms from the beginning of the simulations for 100 s^-1^ and 400 s^-1^, respectively. The t0 guarantees that the measurements concern a steady state independent of initialization artifacts. The numerical experiments show a clear impact of the RBC shape on platelet transport/margination. The more spherized the RBCs the higher the margination.

**Figure 6:**
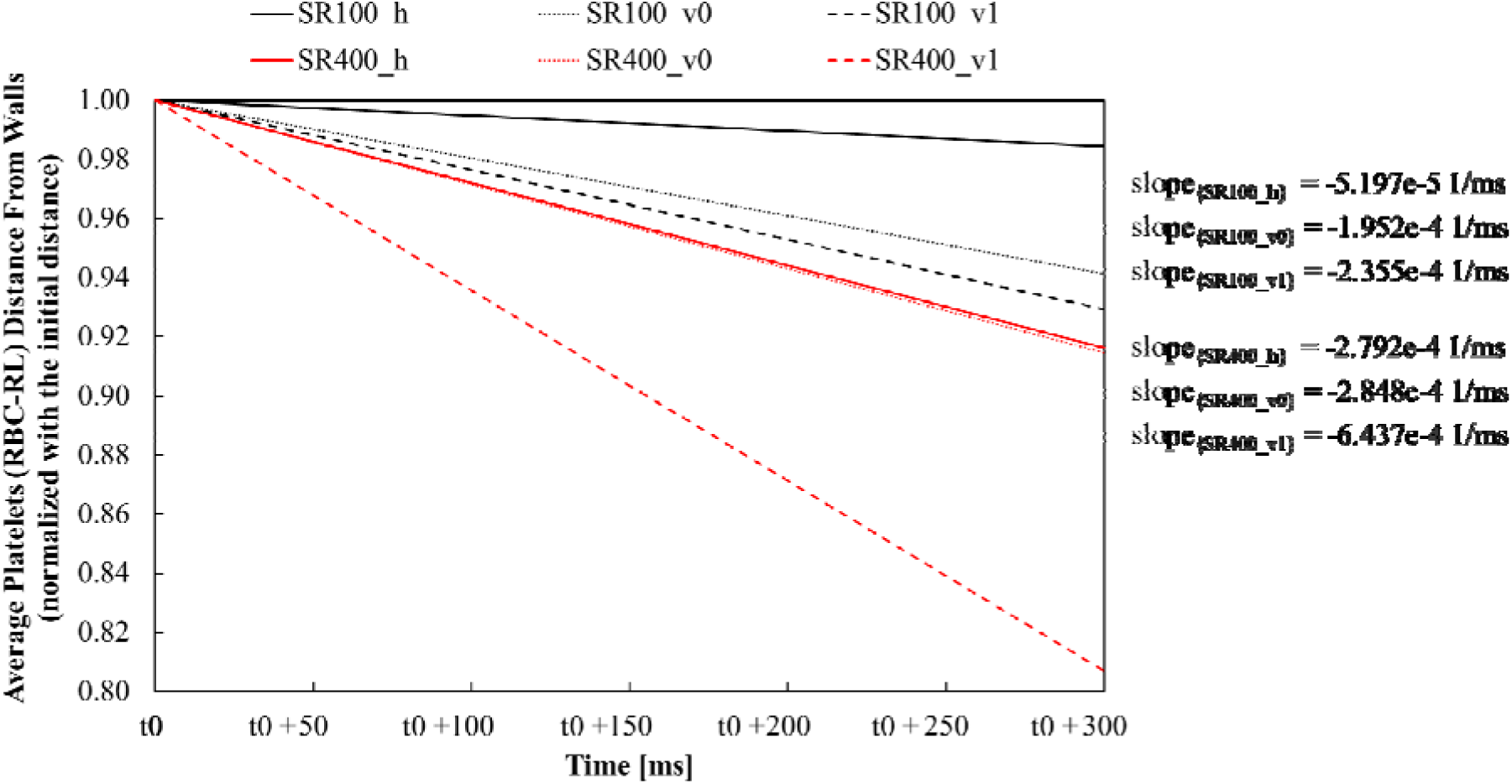
Normalized average distance from walls as extracted from the simulations, by sampling at 10 ms and fitting a line on the extracted data. The behavior of platelets is consistent with the MSD graphs, i.e., the more spherical the RBCs, the faster the platelets are moving towards the walls and the higher the shear rate the faster the transport.

## Discussion

Scientists and clinicians have assumed that RBCs play a passive and unimportant role in thrombus formation. However, nowadays it is clear that RBCs have multiple function and has a clinically significant influence on blood clotting and thrombosis ^19^. Complementary to clinical and experimental studies, computational modeling of thrombosis with a focus on the effects of RBCs has provide some mechanistic explanations ^20^.

In physiological conditions RBCs interactions with the endothelium are minimal. In contrast, in pathological conditions, increased RBCs adhesion to endothelial cells is observed and mediated by several adhesive molecules such as VCAM-1, ICAM-4 etc ^21^. Lysophosphatidylserine has been also pointed out as actor in the adhesion process of RBCs to endothelium ^22^.

Another aspect of involvement of RBCs in thrombus formation is a rheological effect through the platelet margination. In blood flow, RBCs induce a movement of platelets from the center of blood vessel to the wall. So that, they are adjacent to the vessel wall where they can interact to form transitory plugs in case of injury ^23^. Besides, this layer at the surface of endothelium also contain plasma with coagulation factors and white cells.

Another work reported that RBCs can also modulate platelets reactivity directly through the interaction conducted by FasL-FasR interaction ^24^.

Our results show that the sphericity of RBCs increases in COPD stable and exacerbated patients. Previously we observed that RBCs in patients with diabetes, terminal renal failure, sepsis and acute inflammatory state have a more spherical shape than normal subjects evaluated by flow cytometry, the second Pearson coefficient of dissymmetry, (PCD) representing an estimation of the spherical shape ^25^.

How to explain the RBCs shape change in COPD patients?

Gangopadhyay S et al reported that in RBC membrane from COPD patients, phospholipid and cholesterol contents increase. The fatty acid analysis of the individual phospholipids showed preponderance toward saturated fatty acid suggesting a decrease in membrane fluidity that could impair the functions of the RBCs membrane ^26^.

Another work published, using paramagnetic spin label 5-nitroxysterate, reported structural alterations in erythrocytes from patients with COPD. Authors observed an increase of RBCs membrane rigidity ^27^.

Another hypothesis to explain the RBCs spherization is the loss of membrane sialic acid content. It is well known that in chronic or acute inflammation a decrease of sialic acid on RBCs membrane is observed ^9^. Previously, Piagnerelli et al demonstrated a direct effect of neuraminidase on RBCs spherization ^25,28^.

Our experimental results corroborate our hypothesis. Neuraminidase treatment, inducing a spherization of RBCs, modified the adhesion and aggregation of platelets in impact-R. The way in which our experiment is designed does not allow us to know if the observed effect is linked to the decrease in electronegativity of RBCs, associated with the loss of sialic acid, or to an effect simply linked to the form of RBCs.

This is where the digital simulation is complementary to the experimentation.

Indeed, to further support the experimental findings, we deployed the coarse-grained model of the Impact-R device suggested by Chopard et al ^7^. In this work, we replaced the complex dynamics of the bulk with 1D diffusion equations to describe the movement of platelets towards the substrate of the device. These equations define the candidate particles for adhesion/ aggregation and through stochastic rules the authors approximate the deposition process. To cross-check the *in-vitro* experiments and considering an increased transport from the fully resolved 3D simulations (for more spherical RBCs), we increased the diffusion coefficient in the coarse-grained model ^7^ and compared the new values of the aggregate size and number with the in-vitro ones. The coarse-grained model has been validated for shear rate 100 s^-1^ and the amount of increase of the diffusion coefficient was defined by dividing the MSD_h_ (normal RBC shape, 0.1554 μm^2^/ms) with the MSD_v0_ (v0 RBC shape, 0.1943 μm^2^/ms) and MSDv1 (v1 RBC shape, 0.2540 μm^2^/ms). The in-vitro experiments give an increase of about 2.5% of the aggregate size and about 30% of the aggregate number when the shape of RBCs is more spherical. The coarse-grained model, by projecting the RBC shape change on the diffusion coefficient, gives an increase of about 5% of the aggregate size and about 10% of the aggregate number when the shape of RBCs is more spherical. The discrepancy between the results can be explained, given that the coarse-grained stochastic model is based on parameters that are “trained” based on specific patient data. Considering that there is a new set of experiments with potential differences to the ones of Chopard et al ^7^ (different patients), then it is reasonable to consider a qualitative comparison, rather than a strict quantitative one. In conclusion, our results show that the RBCs of COPD patients are more spherical than those of healthy volunteers with an effect more pronounced in ECOPD patients. Experimentally, we observe that the RBCs spherization induces an increase of platelets adhesion-aggregation process. The numerical simulations provide one explanation of experimental results indicating that the RBCs spherization induces an increase of platelets transport to the wall, which in turns gives rise to an increase of the number of aggregates and their size. Additional studies are needed to better understand the possible association between the RBCs effect on the platelets margination and the increased cardiovascular events observed in COPD patients.

## Data Availability

Data are available

